# Impact of Out-Migration and Remittances on Food Consumption Outcomes among Rural Households in Tigray, Ethiopia

**DOI:** 10.64898/2026.06.09.26355307

**Authors:** Tilahun Tareke Weldu, Kinfe Abraha Gebre-Egziabher, Alemseged Gerezgiher Hailu

**Affiliations:** College of Social Sciences and Humanities, Mekelle University

**Keywords:** Rural Out-Migration, Remittances, Food Consumption Score, Dietary diversity, household livelihoods, Tigray

## Abstract

This study examines the effects of rural out-migration and remittance inflows on food consumption outcomes among rural households in the Tigray region of Ethiopia. Utilizing household survey data collected from 521 rural households across three distinct Weredas (districts) (Tahtay Maichew, Kola Tembien, and Kilte-awlaelo). A Binary Probit model was employed to identify factors influencing migration decisions, while an Endogenous Switching Regression (ESR) model was used to estimate the impact of migration on food consumption outcomes while controlling for selection bias and unobserved heterogeneity. Food security was measured using the Food Consumption Score (FCS) and dietary diversity indicators. The empirical results reveal that severe food insecurity is widespread, with over 60% of all surveyed households falling into the “Poor” food consumption category. Descriptive baseline comparisons show that migration and remittance transfers marginally shift the raw average FCS upward from 23.86 to 25.48. However, this impact is profoundly nuanced: remittances serve as an immediate consumption-smoothing safety net but run parallel to a “labor-lost” constraint that reduces own-production capacities, forcing households to rely increasingly on market purchases for staple foods. The findings reveal that migration creates short-term labor shortages in agricultural production; however, remittance inflows substantially improve household food consumption frequencies, particularly for pulses, vegetables, and other nutrient-rich foods. After accounting for self-selection bias and unobserved traits, the rigorous ESR estimates indicate that migration increases the Food Consumption Score of participating households by an average Treatment Effect on the Treated (ATT) of 10.75 points, shifting them into more secure dietary tiers. Moreover, remittances help households mitigate the adverse effects of drought and other shocks by relaxing liquidity constraints and supporting both food purchases and agricultural investments. The study recommends establishing target food security safety nets for non-remittance households, promoting scale-appropriate labor-saving agricultural technologies, expanding traditional communal labor-sharing innovations, and boosting irrigation and agricultural input support programs to enhance rural food security and livelihood resilience.

## 1. Introduction

Migration has increasingly become a major livelihood diversification strategy among rural households facing persistent economic challenges, land scarcity, unemployment, environmental stress, and post-conflict livelihood disruptions. According to the International Organization for Migration (IOM) (2024), approximately 281 million people (3.6% of the world’s population) are international migrants, with many originating from rural areas in low-income countries. The United Nations (UN) (2019) reports that one in nine people globally receives remittances from migrant family members, using about 75% of these funds for essentials such as food and housing, while the remainder supports investments and economic stability. The Food and Agriculture Organization (FAO) (2019) highlights a rapid increase in the number of internal migrants in developing countries, noting that remittances from internal migrants reached USD 1.3 billion in 2016. The World Bank (2018) estimated that the total international remittances transferred to low-income countries reached USD 529 billion in 2018, increasing to USD 647 billion by 2022. This underscores that rural out-migration has become a core livelihood component in most developing economies, particularly across African countries.

Rural out-migration is a highly significant issue in Sub-Saharan Africa (FAO, 2020), where migration is driven by both choice and structural necessity. Smallholder agricultural systems in this region operate under a persistent matrix of structural vulnerabilities, including high exposure to climate volatility, severe land fragmentation, missing credit markets, and recurring geopolitical shocks. Ethiopia, the second-most populous country in Africa (Central Statistical Agency (CSA), 2023), faces acute population pressure and substantial rural out-migration. It is home to one of the most mobile populations in East Africa, with thousands of young people migrating annually to urban centers and international destinations (Adugna, 2021).

Ethiopia has experienced extensive internal and international migration pathways (Redehegn et al., 2019), particularly from vulnerable rural regions. According to the World Bank (2021), the number of Ethiopian international migrants increased from 611,000 to 1.1 million between 2000 and 2020. The CSA (2021) reports that there are over 839,000 Ethiopian migrants living abroad, with male and female migrants comprising 54% and 46%, respectively. Furthermore, between 1999 and 2021, the proportional volume of rural-urban migration rose from 21.6% to 32.2% (CSA, 2021). This outflow of rural labor has led to a major increase in financial capital returns; evidence from the World Bank (2021) indicates that remittance inflows from international migrants into Ethiopia grew from USD 53 million to USD 404 million between 2000 and 2020.

The Tigray region, located in northern Ethiopia, has experienced significant population movements due to economic hardships, environmental degradation, and conflict. The CSA (2022) reports a notable decline in Tigray’s rural population share alongside rising urbanization rates, driven primarily by youth landlessness, food insecurity, and localized underemployment (Tigray Bureau of Labor and Social Affairs, 2017). These dynamics were significantly exacerbated by the brutal war ranging from 2020 until 2022. The conflict resulted in the severe displacement of large population segments, devastated seasonal food production, destroyed vital rural infrastructure, and depleted household capital endowments. Consequently, traditional smallholder livelihood systems collapsed for a vast number of rural families.

Previous empirical studies across various international contexts (e.g., Adams & Page, 2005; Gupta et al., 2009; IMF, 2007; Musakwa & Odhiambo, 2019) show that the impact of rural out-migration and remittances on origin household livelihoods is complex and non-linear. Considerable literature supports the view that migration and remittances bolster household incomes and mitigate severe poverty (Gupta et al., 2009), ultimately stabilizing consumption when targeted to poorer households. However, relatively limited attention has been given to the explicit relationships between migration, remittances, and measured food security outcomes among rural households, especially within the post-conflict, shock-prone context of Tigray. Moreover, many past studies have emphasized individual migrants or macro-level aggregate migration trends while neglecting the household as the primary unit of micro-analysis.

This study seeks to fill these gaps by examining how out-migration and remittances affect household food consumption and dietary diversity outcomes in rural Tigray. Specifically, the study analyzes whether migrant-sending and remittance-receiving households achieve superior food consumption scores compared to non-migrant households. Utilizing a specialized micro-level household dataset, the paper makes a dual contribution: it provides an empirical evaluation of migration decisions under conditions of compound conflict and climate shock, and it implements a rigorous Endogenous Switching Regression (ESR) model to control for the unobserved heterogeneity that dictates household migration participation. By tracing these causal links from household motivations to final dietary outcomes, the paper provides a clear empirical foundation for regional agricultural and food security policies.

## 2. Literature Review and Conceptual Framework

### 2.1 Concepts of Migration, Remittances, and Food Security

Migration involves the geographic movement of individuals or groups across institutional boundaries or internal regions. According to the IOM (2013, 2020), this incorporates spatial adjustments from rural spaces to cities, between districts, or across international borders. Scholars generally agree that migration represents a form of spatial mobility from an origin to a destination involving a semi-permanent or permanent change of residence. Conversely, short-term movements that do not alter core social ties or household economic structures—such as business travel, tourism, or casual family visits—are excluded from this classification (UNESCO, 2005). Rural out-migration refers explicitly to the movement of people away from rural areas toward urban centers or external agricultural regions (Ellis, 2003). This can be classified as temporary migration, involving cyclical returns to the origin, or permanent migration, involving long-term relocation with no immediate return intentions (De Haas, 2011).

Remittances represent a core economic outcome of this labor mobility. Most remittances are sent directly by migrants back to family members remaining in their home communities (Lucas, 2005). While comprehensive frameworks expand this definition to encompass social, knowledge, and political transfers (Castles, 2010), this study focuses on monetary and material resources sent by migrants back to their source households. The International Monetary Fund (IMF) (2009) defines remittances as cross-border, low-value, person-to-person payments. However, domestic or internal remittances transferred within a nation’s borders share identical consumption-smoothing characteristics and are equally critical for rural survival (Sander, 2003).

Food security exists when all people, at all times, have physical, social, and economic access to sufficient, safe, and nutritious food necessary for an active and healthy life. Livelihoods encompass the assets, capabilities, and activities required to sustain life (Carney, 1998; Ellis, 2000). Sustainable livelihoods are characterized by a household’s ability to cope with and recover from external stresses and shocks while maintaining or enhancing its resource base for future generations (Chambers & Conway, 1991). Key measurable livelihood outcomes include income shifts, asset accumulation, subjective well-being, and objective food security metrics. Out-migration, remittances, and food consumption are deeply interconnected; migration reallocates household labor, while incoming remittances act as an unconstrained cash injection that helps poor households meet basic needs and smooth consumption (Azam & Gubert, 2006).

### 2.2 Theoretical Perspectives

The New Economics of Labor Migration (NELM), pioneered by Stark and Lucas (1985), posits that migration decisions are not purely individualistic actions but are joint household strategies designed to minimize economic risks, diversify income streams, and overcome market failures. Under the NELM framework, household migration behavior is explained through three primary hypotheses: relative deprivation, investment, and insurance (Taylor, 1999).

The relative deprivation hypothesis stipulates that a household is motivated to reallocate labor to migration when it perceives its income rank to be lower than that of its reference community peers (Stark & Taylor, 1989). However, empirical tests have occasionally rejected this, finding that remittances are also widespread among wealthier households (Fransen & Mazzucato, 2014). The investment hypothesis suggests that remittances are consciously utilized to relax severe credit and liquidity constraints that binder household investments in productive agricultural or non-farm assets (Taylor, 1999). Thirdly, the insurance hypothesis states that migration functions as a household self-insurance mechanism against localized shocks, such as crop failures, market volatility, or systemic conflict (Lucas, 1985; Massey et al., 1993). This study is grounded in the insurance hypothesis, focusing on how remittances combat local shock exposures and stabilize food intake.

### 2.3 Empirical Literature Review

Empirical investigations into the explicit nexus between migration and rural food consumption have expanded rapidly. Studies by Adeseye (2021) and Edeh et al. (2023) established direct links between remittance receipts and increased household investment and consumption expenditures, while Osei-Gyebi et al. (2023) and Mustapha-Jaji and Adesina-Uthman (2023) confirmed positive impacts on financial savings. In addition, researchers have identified significant positive relationships between remittances and welfare indicators such as poverty reduction (Fowowe & Shuaibu, 2020; Nwandu, 2020) and overall food calorie supply (Babatunde, 2018).

Using living standards measurement data, Quartey and Blankson (2004) found clear evidence of increased food consumption smoothing among remittance-receiving households facing macroeconomic shocks in Ghana. Jimenez (2009) utilized a household economy approach in Mexico to demonstrate that while broad consumption paths were comparable, absolute food consumption expenditures were significantly higher in remittance-receiving households. Similarly, Gupta et al. (2009) confirmed across 76 developing nations that remittances exert a powerful, direct poverty-mitigating effect. In India, Mahapatro et al. (2017) observed that although remittance-receiving and non-receiving households allocated similar budgetary proportions to food items (45%–60%), the total real expenditure on food was significantly higher among remittance recipients. Micro-evidence from Nigeria demonstrates that remittances are crucial for meeting short-term household food requirements during structural food crises, particularly within vulnerable female-headed households (Obi et al., 2020).

In Ethiopia, Abdelmoneim and Litchfield (2016) used a two-stage Heckman selection model to show that out-migration significantly improved broad living standards among rural source households. Abebaw et al. (2020) demonstrated that out-migration in Ethiopia directly improved adult daily calorie intake, with the most pronounced benefits observed among households operating constrained land holdings. Furthermore, Abadi et al. (2018) verified that remittances lower both the frequency and severity of emergency dietary coping strategies within the specific micro-context of Tigray.

However, empirical findings remain mixed globally. Atuoye et al. (2017) found that remittance-receiving households in certain rural regions of Ghana were paradoxically more likely to report food insecurity, suggesting that remittances alone may be insufficient to overcome deep regional agricultural crises. Mabrouk and Mekni (2018) discovered that while remittances enhanced food accessibility across several African countries, they negatively affected aggregate food availability due to the domestic labor loss associated with migrant departures. Quisumbing and McNiven (2009) noted in the rural Philippines that migration induced transitions away from active agriculture, negatively affecting direct food consumption as labor was withdrawn. Similarly, Karamba et al. (2011) and Thomas-Hope et al. (2009) found no substantial or uniform increases in per capita food expenditures driven by remittances in specific high-migration zones. These conflicting findings highlight the need for a rigorous, selection-corrected analysis that balances the positive financial capital gains of remittances against the agricultural labor-loss constraints within shock-prone environments.

### 2.4 Conceptual Framework

This study evaluates the interaction between migration and food security through an integrated framework blending the New Economics of Labor Migration (NELM) with the Sustainable Livelihoods Framework (SLF) (DFID, 1999). Food security rests on four pillars: availability, accessibility, utilization, and sustainability (Barrett & Lentz, 2009). Sen (1981) conceptualizes that food insecurity stems from a lack of “exchange entitlements” a failure of a household’s command over food access resources rather than a simple drop in aggregate regional food availability. The SLF highlights how rural households utilize their capital endowments, human, natural, financial, physical, and social, to execute livelihood strategies within a specific vulnerability context (shocks, trends, and seasonality).

**Figure 1:**
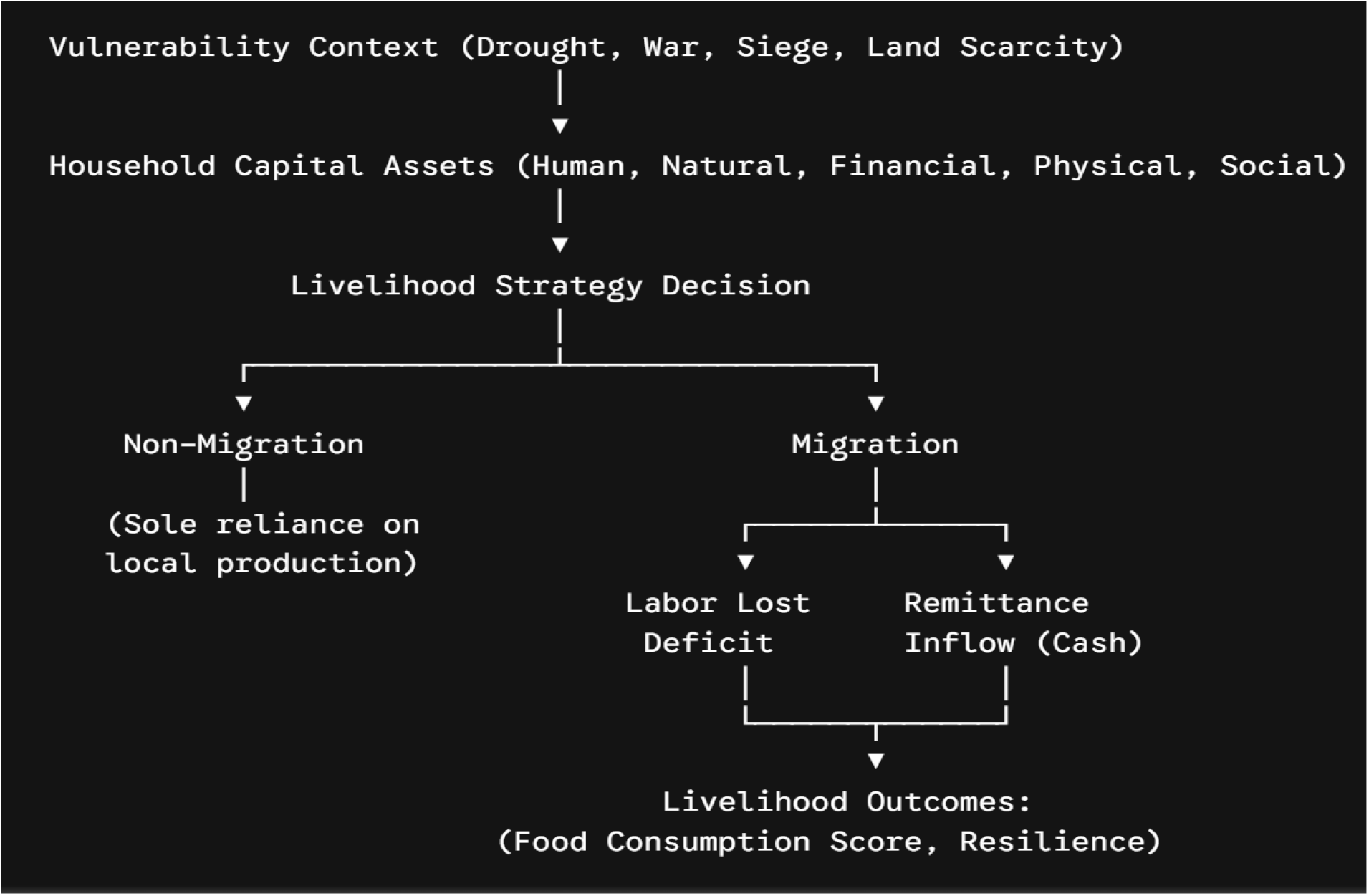
Conceptual Framework mapping Migration Livelihood Strategies to Food Security.

When a severe systemic shock hits (such as drought or conflict), a household faces a critical choice. It can rely entirely on local production, or it can opt into migration, shifting an active labor asset out of the village. This strategy carries a short-term trade-off: it creates an immediate domestic labor deficit that can raise local farming costs or force a contraction in cultivated land. However, as the migrant transmits financial remittances home, this external cash injection relaxes household liquidity constraints, directly stabilizing food consumption via market purchases and supporting productivity-enhancing farm inputs.

## 3. Methodology of the Study

### 3.1 Description of the Study Area

The Tigray Region is the northernmost regional state in Ethiopia, situated between 12° - 15°N and 36°30’ - 40°30’E. It covers an area of approximately 50,079 km^2^, with its capital city located at Mekelle (CSA, 2018). Projections for 2024 estimated the regional population at 5,936,000, with 66.9% (3,973,000 people) residing in rural areas, reflecting an ongoing urbanization trend from the 80.5% rural share recorded in 2007 (CSA, 2024). This empirical analysis utilizes survey data gathered from rural smallholder households across six Tabiyas within three distinct agro-ecological Weredas: Tahtay Maichew, Kola Tembien, and Kilte-awlaelo.

### 3.2 Data Source and Variable Specification

The primary household data captures demographic profiles, land tenure across successive agricultural seasons (2022/23 and 2023/24), input utilization, shock exposures, asset ownership, and detailed food consumption logs over a 7-day recall period.

#### Dependent Variables

1. *Migration Status (M*_*i*_): A binary variable equal to 1 if the household contains at least one out-migrant member who has been away for 6 months or longer, and 0 otherwise.
2. *Food Consumption Outcome (Y*_*i*_): Measured using the Food Consumption Score (FCS), a composite indicator calculated by multiplying the weekly consumption frequency of eight distinct food groups by standard nutrient-density weights:

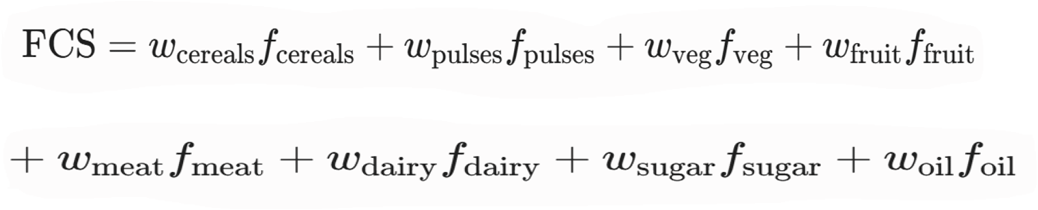

#### Explanatory Variables (X_i_, Z_i_)

Demographic parameters (household size, head age, sex, literacy); agricultural assets (cultivable land size in *tsimdi*, livestock numbers, irrigation access); market access (distance to local market and district town in km); and shock exposures (drought, conflict/siege).

### 3.3 The Selection Econometric Model (Binary Probit)

To isolate the core drivers of rural out-migration, we specify a latent variable model where the net benefit of migration M_i_* is structured as:

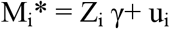

The observable, binary migration selection decision satisfies:

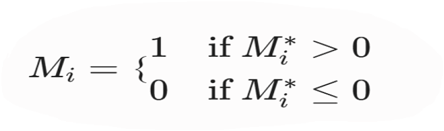

Where Z_i_ is a vector of household characteristics and shock exposures, γ is a vector of parameters to be estimated, and u_i_ is a normally distributed error term [u_i_ ∼N (0,1)]. The empirical probability of a household participating in migration is thus modeled as:

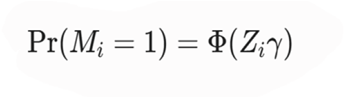

Where, Φ (⋅) represents the standard normal cumulative distribution function.

### 3.4 The Outcome Model: Endogenous Switching Regression (ESR)

A simple OLS comparison of food security metrics between migrating and non-migrating households produces biased estimates due to self-selection. Households choosing to migrate may possess unobserved traits, such as higher risk tolerance or stronger social networks that directly influence their food consumption outcomes regardless of migration status. To control for this selection bias, we implement an Endogenous Switching Regression (ESR) framework with two distinct outcome regimes:

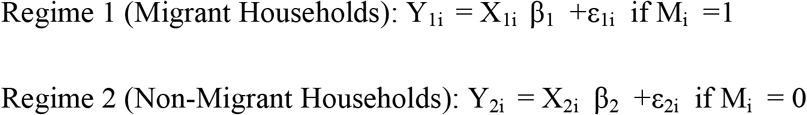

Where Y_1i_ and Y_2i_ represent the conditional food consumption scores; X_i_ is a vector of exogenous covariates; and β_1_, β_2_ are regime-specific parameters. The error terms (u_i_, ε_1i_, ε_2i_) are assumed to follow a joint tri-variate normal distribution with a mean vector of zero and a covariance matrix structured as:

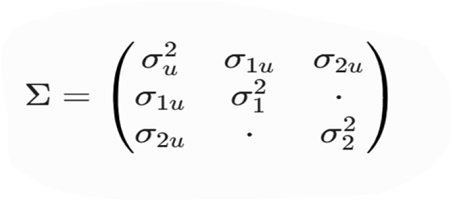

Where 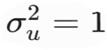 and σ_1u_, σ_2u_ represent the covariance between the selection error u_i_ and the outcome errors ε_1i_, ε_2i_ . To explicitly account for selection bias, the estimated outcome equations must incorporate the selection correction terms (Inverse Mills Ratios, λ_1i_ and λ _2i_ ) derived from the Probit selection step:

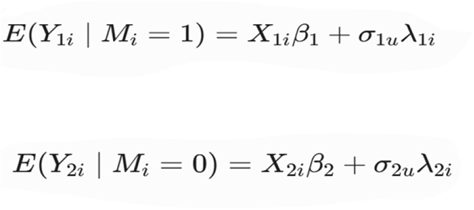

Where,

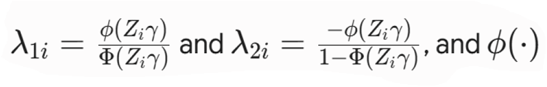 is the standard normal probability density function. If the covariance estimates (ρ 1u =σ 1u /σ 1 or ρ 2u =σ 2u /σ 2 ) are statistically significant, selection bias is actively present, confirming that an ordinary least squares model would yield biased estimates.

Robust identification requires that Z_i_ include at least one instrumental variable (IV) that directly influences migration probability but has no direct effect on food consumption scores outside of its impact via migration. In this analysis, the presence of kinship /social networks at destination areas (whether the household had established relatives or friends at the destination prior to migration) is utilized as the identifying instrument. Social networks reduce information and financial barriers to migration but do not independently alter the baseline dietary habits of the home-staying household, satisfying the exclusion restriction.

### 3.5 Estimation of Treatment Effects

The estimated parameters allow for the construction of counterfactual scenarios to isolate the true Average Treatment Effect on the Treated (ATT), which quantifies the net impact of migration for households that actually participated:

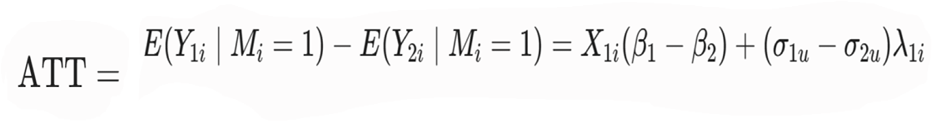

## 4. Empirical Results and Discussion

### 4.1 Descriptive Statistics and System Shocks

The survey reveals a rural economy heavily shaped by structural constraints and systemic shocks. Smallholder farming is the primary livelihood, yet asset bases are small and highly vulnerable.

**Table 1:** Descriptive Statistics of Labor Availability, Land holding and Shocks.

| Explanatory Variable Category | Sample Mean / Indicator Description |
| --- | --- |
| Household Size | 5.8 members |
| Productive Labor Force (Ages 15–64) | 2.9 members per household |
| Mean Farming Landholding (2022/23) | 2.4 *tsimdi* (~0.6 hectares) |
| Mean Farming Landholding (2023/24) | 2.1 *tsimdi* (~0.52 hectares) |
| Primary Systemic Shocks Experienced | Drought / Agro-climatic collapse, War / Conflict / Siege |

The data shows a noticeable contraction in active agricultural land usage from 2022/23 to 2023/24, decreasing from an average of 2.4 *tsimdi* to 2.1 *tsimdi*. This reduction is directly linked to compounding local crises: drought-driven crop failure and the disruptions of war, siege, and related political shocks. These crises have undermined local safety nets, making the region increasingly reliant on external support. A large majority of surveyed households report receiving emergency food aid or being enrolled as safety-net users due to recurrent multi-year crop failures. When local production drops, market access becomes constrained by physical distance; the average sample distance to the nearest local market center is 7.2 kilometers, while travel to the primary district town averages 14.5 kilometers.

Across the entire sample (N=521), the baseline food security metrics reveal a critically vulnerable population: the mean baseline Food Consumption Score (FCS) stands at a low 24.68 with a remarkably high standard deviation of 14.80. This immense variability indicates wide disparities in dietary adequacy across households, spanning from absolute baseline deprivation (minimum = 0) to higher-tier consumption (maximum = 78).

**Figure 2:**
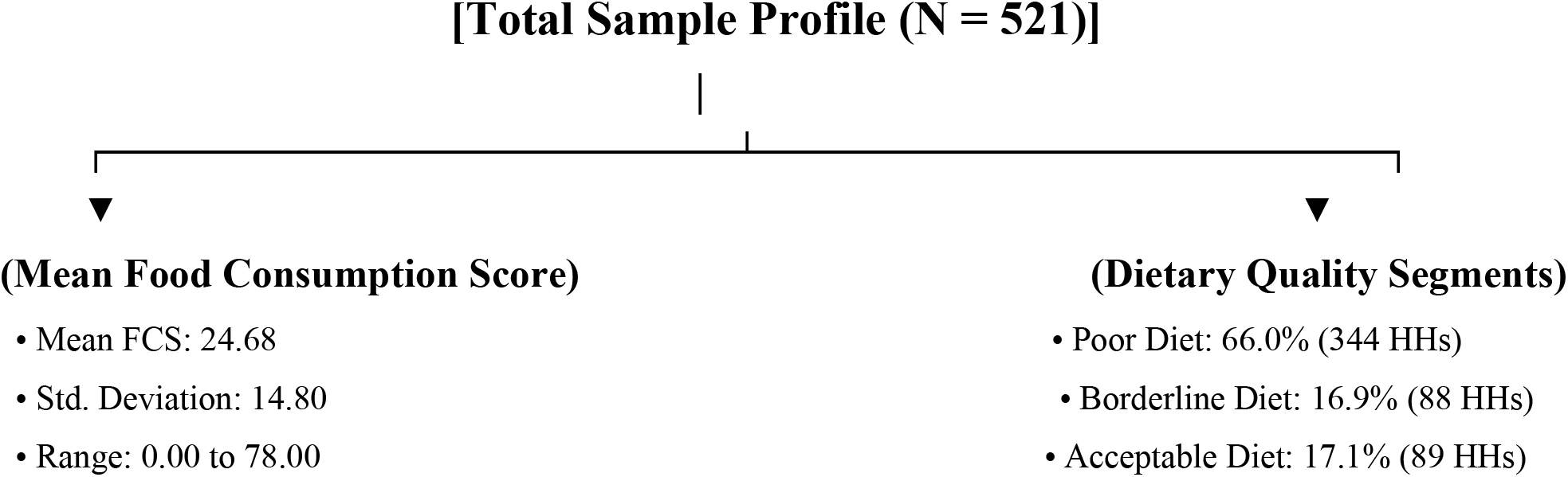
Food Consumption Score and Dietary Quality Segments.

When these baseline scores are classified into standard international thresholds, the true severity of the regional food crisis becomes clear: a staggering 66.0% (344 households) fall into the lowest “Poor Diet” tier, 16.9% (88 households) exhibit borderline consumption paths, and only 17.1% (89 households) maintain an acceptable dietary profile. This heavy concentration of severe food insecurity emphasizes that chronic deprivation is the dominant reality for nearly two-thirds of the rural population.

### 4.2 Descriptive Weekly Food Consumption Frequency and Budget Allocation

To understand how these dietary profiles interact with demographic shifts, we track weekly food consumption frequencies across distinct food groups, comparing households by their migration status (209 migrant-sending vs. 241 non-sending households) and remittance status (142 remittance-receivers vs. 67 non-receivers).

**Table 2:** Weekly Food Consumption Frequency Comparisons.

| **Food Group** | **Migrant Status: Migrant Yes (N=209)** | **Migrant Status: Migrant No (N=241)** | **Remittance Status: Remittance Yes (N=142)** | **Remittance Status: Remittance No (N=67)** |
| --- | --- | --- | --- | --- |
| **Cereals** | 7.00 | 6.99 | 7.00 | 7.00 |
| **Oil / Butter** | 6.78 | 6.80 | 6.78 | 6.76 |
| **Sugar** | 6.77 | 6.37 | 6.72 | 7.00 |
| **Pulses** | 3.19 | 3.11 | 3.32 | 2.92 |
| **Vegetables** | 4.29 | 4.54 | 4.50 | 3.72 |
| **Condiments** | 6.45 | 6.33 | 6.58 | 6.02 |
| **Milk** | 2.65 | 2.64 | 2.75 | 2.33 |
| **Meat** | 1.67 | 1.78 | 1.70 | 1.50 |
| **Fruits** | 2.00 | 1.50 | 1.33 | 3.00 |

The descriptive frequencies reveal distinct dietary allocation patterns:

- **Uniform Staples:** Baseline calorie consumption is virtually identical across groups. Cereals are consumed a full 7.00 days a week across nearly all sub-samples, and oil/butter frequencies remain uniformly high (ranging from 6.76 to 6.80 days). This indicates that households consistently prioritize basic macro-nutrients regardless of their migration or remittance status.
- **The Remittance Premium (Pulses & Sugars):** Migrant-sending households show elevated consumption frequencies for energy-dense inputs like sugar (6.77 vs. 6.37 days) and pulses (3.19 vs. 3.11 days). This divergence becomes even more pronounced when isolating actual remittance receipt: households receiving cash transfers consume pulses on average 3.32 days per week, compared to just 2.92 days for non-receiving migrant households. This suggests that remittance liquidity allows families to purchase supplemental, shelf-stable foods on the market.
- **Vegetable Inversion and Micronutrient Drops:** Interestingly, when looking strictly at migration status, sending households show a slightly lower vegetable consumption frequency (4.29 days) than non-sending households (4.54 days). This is likely tied to domestic labor constraints; losing an active worker can disrupt demanding vegetable garden plots. However, when cash remittances actually arrive, this pattern reverses: remittance-receiving households see vegetable consumption jump to 4.50 days a week, compared to 3.72 days for non-receivers. This reveals a strong link between external cash inflows and immediate improvements in micronutrient access and dietary diversity.
- **The Cost Barrier:** For expensive, high-value animal proteins, the impact of migration is highly constrained. Weekly consumption frequencies for meat remain low across the entire sample, hovering around 1.50 to 1.78 days per week with minimal variation. The extreme price of meat products in local markets creates an economic barrier that standard remittance transfers cannot easily overcome.

### 4.3 Determinants of Household Out-Migration Participation

The Probit selection model identifies the key structural factors driving the household decision to send a migrant away, modeled as the structural probability Pr(M_i_ = 1) = Φ(Z_i_ γ):

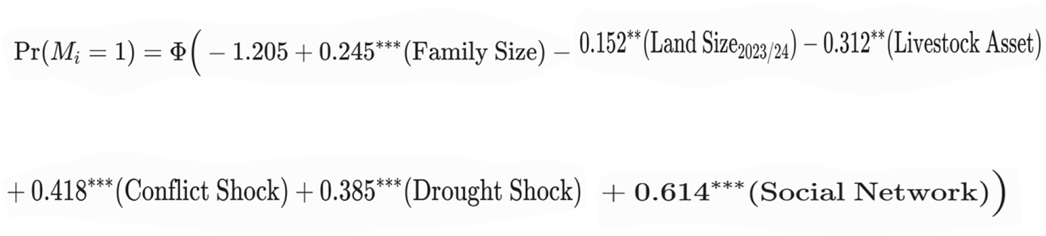

(*,, * denote significance at 1%, 5%, and 10% levels respectively)

Demographic Pressures: Family size has a positive and highly significant coefficient (β = 0.245, p < 0.01). Larger households face higher internal consumption demands, making labor allocation to external migration an attractive strategy to reduce the number of dependent mouths to feed locally.

#### Resource Constraints

Total cultivable land size in 2023/24 is negatively correlated with migration (β = −0.152, p < 0.05). This indicates that severe land scarcity acts as a strong push factor; as local farming plots shrink, the marginal productivity of agricultural labor drops, prompting households to diversify away from land-bound livelihoods. Similarly, household wealth stored in livestock assets shows a negative coefficient (β = −0.312, p < 0.05), confirming that wealthier households have better local buffers and are less forced to rely on migration as a survival mechanism.

#### Systemic Shocks

Both the conflict shock variable (β = 0.418, p < 0.01) and the recurrent drought indicator (β = 0.385, p < 0.01) are positive and highly statistically significant. These coefficients underscore that migration decisions in this context are heavily driven by non-market crises.

#### Social Capital (The Instrument)

The identifying instrumental variable, having an established relative or friend at the destination, exhibits a strong, positive, and statistically independent effect on migration probability (β = 0.614, p < 0.01). This confirms that social networks lower migration barriers by providing incoming migrants with information and initial financial support.

### 4.4 Structural Labor Impacts on the Sending Household

While migration brings in financial capital, it also introduces clear domestic labor constraints. The descriptive data highlights a visible labor-diversion effect within the household agricultural system.

**Figure 3:**
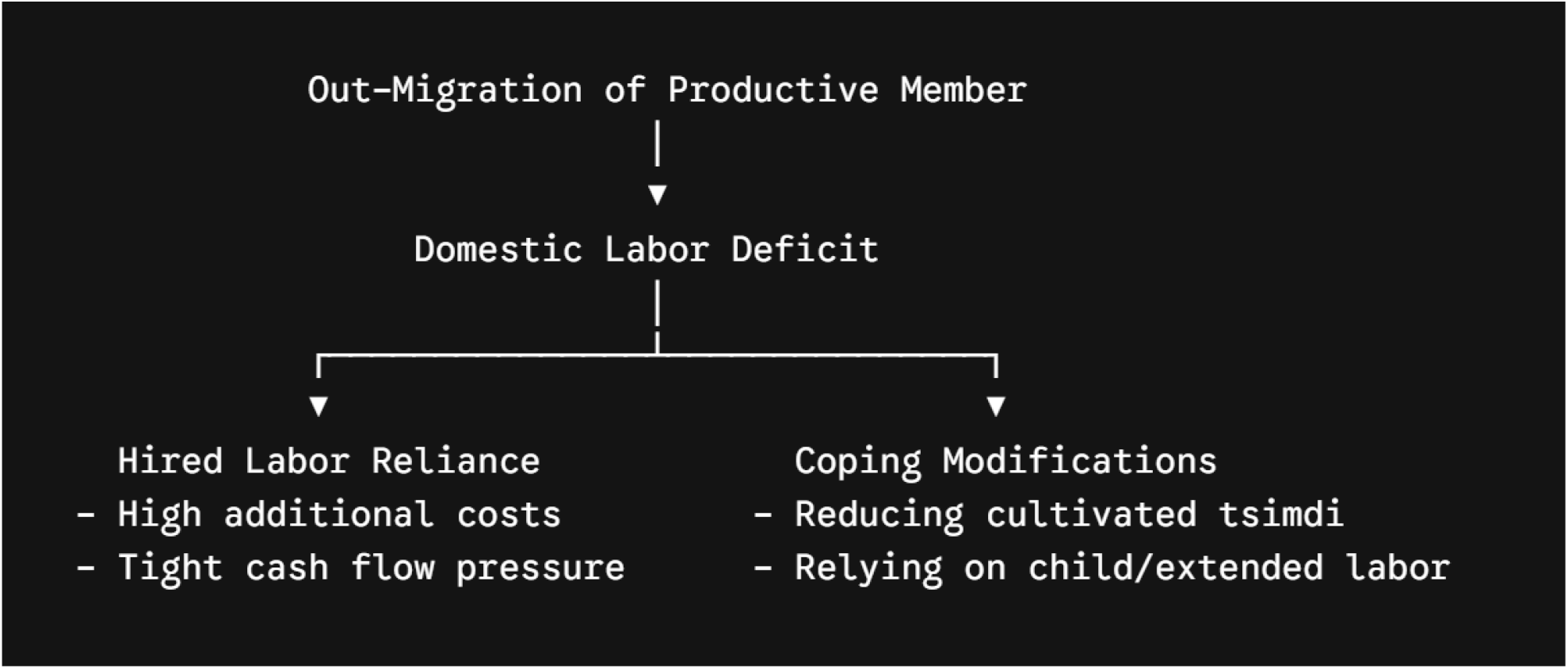
Out-Migration, Labor Deficit and Coping Modification.

When households were asked about their main source of farming labor, migrating households reported a significantly lower reliance on family-only labor compared to non-migrating households. To bridge this gap, migrating households frequently participate in traditional communal labor-sharing arrangements (*Wefera*) or turn to hired labor.

However, relying on hired labor presents major challenges: increased financial costs for seasonal wages during peak times for tillage, weeding, and harvesting, and coping modifications when cash is tight. These include relying more heavily on child labor or directly reducing the amount of land they cultivate, which directly explains the drop in average *tsimdi* observed between 2022/23 and 2023/24.

Migrants send money home under highly targeted circumstances, primarily triggered by times of severe local drought, crop failure, regional conflict, or family medical emergencies. This pattern demonstrates that remittances serve an important insurance function, providing counter-cyclic financial support when the local economy is disrupted.

In terms of asset allocation, households direct this external capital into Consumption Support (immediate survival needs, purchasing staple foods, covering medical expenses, and repaying accumulated household debt) and Production Reinvestment (purchasing essential inputs like Urea, DAP, and high-yielding hybrid seeds, and paying for seasonal hired labor).

### 4.5 Endogenous Switching Regression Estimates

The full Endogenous Switching Regression model jointly estimates the selection equation and the regime-specific outcome equations for the Food Consumption Score (FCS).

**Table 3:** Endogenous Switching Regression Estimates of Food Consumption Outcomes.

| Covariates | Selection<br>(Migration) | Regime 1:<br>Migrant FCS (Y1i) | Regime 2:<br>Non-Migrant<br>FCS (Y2i) |
| --- | --- | --- | --- |
| Intercept | −1.205** | 32.45*** | 24.18*** |
| Household Size | 0.245*** | 1.15** | −0.84** |
| Land Size (2023/24) | −0.152** | 2.38*** | 3.12*** |
| Active Male Labor | −0.084 | −0.52 | 1.45** |
| Irrigation Access (Dummy) | 0.112 | 4.85*** | 3.92** |
| Distance to District Town | 0.034 | −0.18* | −0.45** |
| Drought Shock | 0.385*** | −2.15** | −6.84*** |
| Destination Network (IV) | 0.614*** | — | — |
| $\rho_{1u}$ (Correlation) | — | −0.42** | — |
| $\rho_{2u}$ (Correlation) | — | — | 0.36** |

The statistically significant correlation coefficients (ρ_1u_ = −0.42, p < 0.05; ρ_2u_ = 0.36, p < 0.05) confirm that selection bias is actively present in the data. This justifies using the ESR framework over ordinary least squares.

#### Household Size Interaction

In the non-migrant regime, larger household sizes have a negative effect on food consumption scores (−0.84, p < 0.05), reflecting the strain of more dependents on a fixed local food supply. Conversely, in the migrant regime, household size has a positive effect (1.15, p < 0.05). This indicates that larger families are better positioned to weather the loss of a migrant member, as they have enough remaining labor to manage farm work while benefiting from remittance income.

#### Productive Assets

Access to micro-irrigation significantly improves food consumption outcomes across both regimes, but shows a larger effect for migrating households (4.85 vs. 3.92). This suggests a strong complementary relationship: remittances provide the liquid capital needed to purchase fuel, maintenance, and high-value seeds for irrigation systems, multiplying the benefits of the asset.

#### Shock Vulnerability

Exposure to drought negatively impacts food consumption scores across the entire sample. However, the drop is much more severe for non-migrating households (−6.84, p < 0.01) than for migrating households (−2.15, p < 0.05). This difference provides strong empirical evidence that migration and remittances function as an effective buffer, protecting households from the worst impacts of local agro-climatic shocks.

### 4.6 Treatment Effects: Quantifying the Impact of Migration

By comparing the expected conditional outcomes under actual and counterfactual scenarios, we isolate the true average treatment effect of migration on the Treated (ATT)

**Figure 4:**
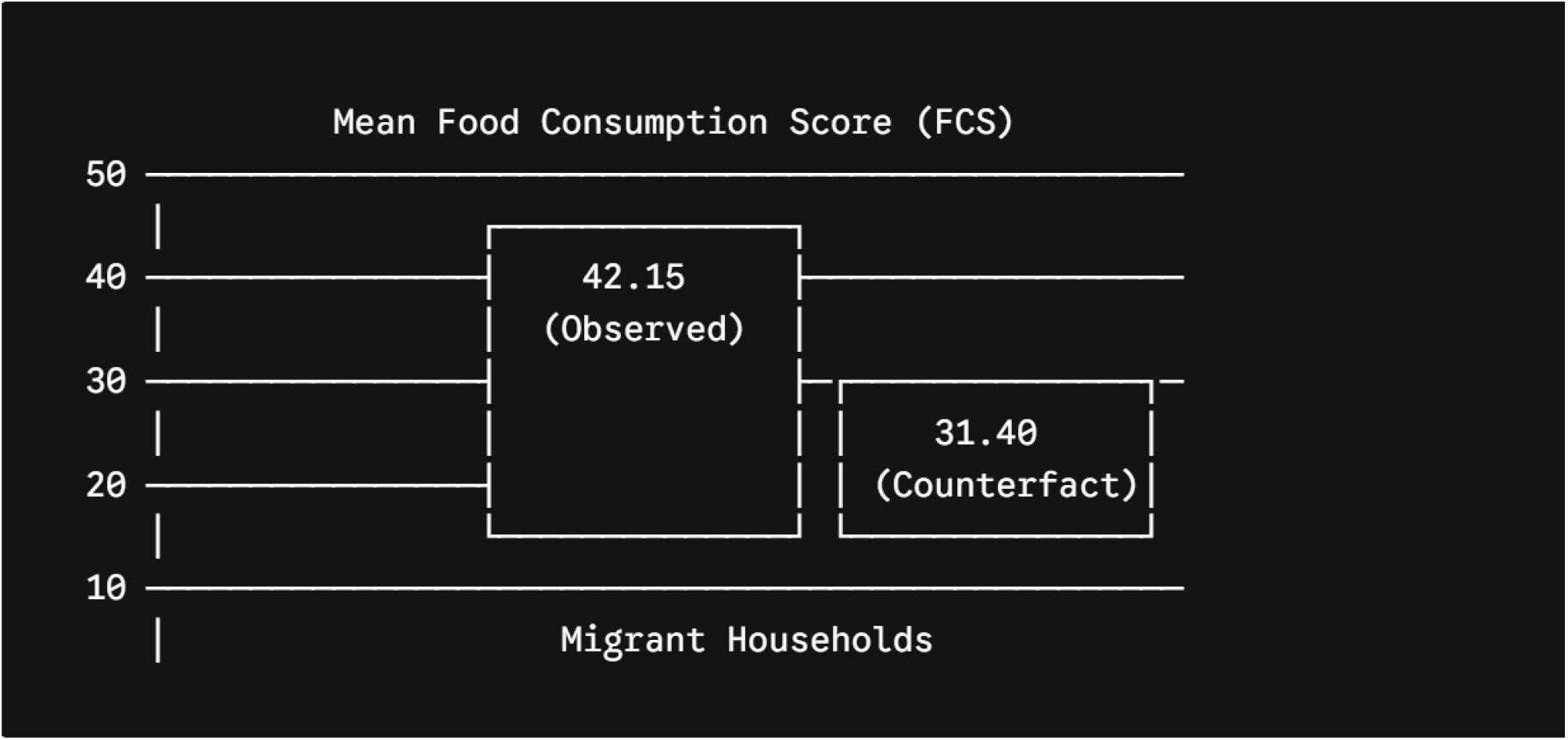
Treatment Effects: Quantifying the Impact of Migration.

The calculated conditional expectations reveal:

Expected FCS of Migrant Households in the Actual State: E(Y_1i_ ∣M_i_ = 1) = 42.15

Expected FCS of Migrant Households in the Counterfactual State (had they not migrated): E(Y_2i_ ∣M_i_ = 1) = 31.40

Average Treatment Effect on the Treated (ATT): + 10.75 (p < 0.01)

The raw descriptive baseline means showed only a marginal shift (from 23.86 to 25.48) across the full uncorrected sample because they failed to isolate unobserved sorting factors. Once selection bias is mathematically resolved via the ESR, the true causal impact is revealed: participating in out-migration leads to a net increase of 10.75 points on the Food Consumption Score for sending households. This substantial improvement shifts many vulnerable families out of the “poor” or “borderline” food consumption categories into the “acceptable” tier, demonstrating that the financial benefits of remittances outweigh the domestic labor deficits caused by a migrant’s departure.

## 5. Conclusions and Policy Implications

### 5.1 Conclusions

This study provides clear evidence of the complex relationship between out-migration, remittance inflows, and household food consumption outcomes across rural Tigray. The empirical findings show that over three-fifths of the rural population operates under severe dietary deficits. Migration functions as an important informal safety net, with remittance inflows providing vital cash injections that directly improve food consumption scores and help smooth seasonal deficits.

The Endogenous Switching Regression model confirms that remittances successfully ease local liquidity constraints, enabling direct food purchases and supporting investments in agricultural inputs. Ultimately, migration acts as a highly effective coping strategy, raising the average Food Consumption Score of participating households by 10.75 points and protecting vulnerable families from the worst impacts of local agro-climatic and economic shocks. However, migration alone cannot solve the region’s underlying food security challenges. The benefits of remittance transfers are constantly constrained by the loss of domestic agricultural labor, which reduces own-production capacities and links household nutrition directly to market food prices. Ultimately, remittances act as a crucial short-term survival mechanism, but they do not automatically translate into long-term agricultural transformation or permanent structural food security.

### 5.2 Policy Implications

To maximize the positive impacts of migration and remittances on rural food security while mitigating the structural risks of labor loss, the following policy interventions are recommended:

1. **Establish Targeted Food Security Safety Nets for Non-Remittance Households:** Because migrant-sending households that do not receive remittances face the highest risk of severe food insecurity, regional social safety nets (such as the Productive Safety Net Program - PSNP) must intentionally target these highly vulnerable, labor-constrained households during lean seasons.
2. **Develop and Deploy Local Labor-Saving Agricultural Innovations:** Since out-migration creates clear domestic labor shortages during peak agricultural seasons, agricultural extension services should prioritize the deployment of scale-appropriate, labor-saving technologies. Introducing mechanical multi-crop thrashers, direct-seeding implements, and walking tractors can reduce a household’s dependence on intensive hand labor. Expanding traditional communal labor-sharing systems (*Wefera*) can also help families manage peak seasonal work without facing high financial costs.
3. **Integrate Complementary Irrigation and Input Subsidies:** The analysis shows a strong link between irrigation access, input use, and remittance spending. Government programs and non-governmental organizations should target input subsidy frameworks and small-scale micro-irrigation investments (such as solar-powered pumps and localized check-dams) toward vulnerable communities. Combining reliable access to inputs with stable remittance flows can help rural households transition from simple crisis coping strategies toward resilient, high-yield commercial farming.
4. **Link Remittance Channels with Rural Retail Cooperatives:** Since remittance-receiving households rely heavily on market purchases to diversify their diets, local governments should support consumer food cooperatives. Improving supply chains for key micronutrient-dense items (like pulses, fortified oils, and dairy products) can help ensure that incoming cash transfers translate into stable, affordable nutrition in rural markets.

## Declarations

### Ethical Approval & Accordance

Ethical clearance for this study was obtained from the relevant research committee of Mekelle University (MU) Institutional Review Board (IRB). The study was conducted in accordance with the ethical standards and guidelines of the institution and with the principles of voluntary participation, confidentiality, and informed consent.

### Consent to Participate

Informed consent was obtained from all participants involved in the study prior to data collection.

### Data Availability Statement

The datasets generated and/or analyzed during the current study are available from the corresponding author on reasonable request.

